# Gastric Alimetry^®^ testing and healthcare economic analysis in nausea and vomiting syndromes

**DOI:** 10.1101/2023.09.07.23295185

**Authors:** William Xu, Lucy Williams, Gabrielle Sebaratnam, Chris Varghese, Chris Cedarwall, Charlotte Daker, Celia Keane

## Abstract

**Background:** Chronic nausea and vomiting syndromes (CNVS), gastroparesis and functional dyspepsia (FD) are complex disorders. Body Surface Gastric Mapping (BSGM), a new test of gastric function, using Gastric Alimetry^Ⓡ^(Alimetry, New Zealand) may be useful for de-escalating healthcare utilisation. This study aimed to define healthcare costs and estimate health economic impacts of implementing this test in patients with chronic gastroduodenal symptoms.

**Methods:** Consecutive patients at a tertiary referral centre evaluated with Gastric Alimetry were included. Frequency and cost data relating to medical investigations, hospital, and outpatient presentations were evaluated. Costs of healthcare utilisation were calculated, and the potential cost savings of implementing Gastric Alimetry within a diagnostic decision-tree model were estimated.

**Results:** Overall, 31 consecutive patients (mean age 36.1 years; 83.9% female; predominant symptoms: nausea [83.9%], pain [61.3%], vomiting [67.7%], bloating [35.5%]) completed Gastric Alimetry testing. Repeat gastroscopy and abdominal CT rates were 29% (8/28) and 85% (11/13) respectively. Gastric Alimetry testing identified spectral abnormalities in 45.2% of patients, and symptom profiling classified a further 29.1% of patients. Median annualised cost difference after test introduction was NZ$-12,032. Estimated reductions in investigation-related costs when incorporating Gastric Alimetry into the diagnostic workflow model were approximately NZ$1,500 per patient.

**Conclusions:** Healthcare utilisation and confirmatory testing rates remain high in nausea and vomiting syndromes. This study presents real-world data, together with a decision tree analysis, showing Gastric Alimetry can streamline clinical care pathways, resulting in reduced healthcare utilisation and cost.

## Introduction

Chronic nausea and vomiting syndrome (CNVS) and gastroparesis are debilitating disorders that are difficult to diagnose and manage.^1^ There is a combined global prevalence of ∼1%.^2^ Although differentiated by the presence or absence of delayed gastric emptying, this distinction is controversial as gastric emptying is variable over time, correlates weakly with symptoms, and may not reflect the primary disease mechanism.^3–5^ An umbrella term of nausea and vomiting syndromes (NVS) may be used.^6^ In addition, NVS overlaps with functional dyspepsia (FD), sharing common pathophysiological features including neuromuscular dysfunction and gut-brain dysregulation.^1,5^

Patients with NVS have a disproportionately high healthcare utilisation.^7,8^ Factors contributing to this include complex diagnostic pathways, high rates of hospitalisation, trial and error therapies, implementation of nutritional support, and invasive therapies.^7,9,10^ When diagnostic uncertainty arises, this commonly leads to a ‘process of exclusion’, which can take several years, after which patients still frequently face high rates of repeated confirmatory tests in practice. Such tests often do not contribute further to management but incur further costs.^7,8,11^

Body Surface Gastric Mapping (BSGM) using Gastric Alimetry is a new test of gastric function, offering novel insights into gastric disease pathophysiology.^12,13^ This test aids in phenotyping patients into likely disorders related to the gastric neuromuscular dysfunction versus gut-brain dysregulation, as well as assisting in identifying other contributing factors including vagal, sensorimotor or post-surgical mechanisms.^14,15^ Early cohort studies have indicated that Gastric Alimetry can significantly reduce costs by decreasing the need for investigations, admissions and nutritional support in NVS patients,^16–18^ however, more formal healthcare utilisation evaluations are required.

The aims of this study were therefore to define the healthcare utilisation and costs of patients with NVS, and to estimate the potential health economic impact of implementing Gastric Alimetry testing. Real-world data was included to inform the analysis from a cohort of patients with chronic NVS undergoing work-up including with Gastric Alimetry in Auckland, New Zealand. The hypothesis was that Gastric Alimetry would reduce diagnostic costs for patients with chronic nausea and vomiting symptoms, when seen initially by a gastroenterologist, through consideration of additional investigation costs to the healthcare system, in comparison to current standard of care.

## Methods

Ethics approval was granted by the Auckland Health Research Ethics Committee (AH1352).

### Patient selection

Adult consecutive patients (aged ≥ 16 years) presenting with symptoms of nausea and/or vomiting, as well as overlapping features of FD evaluated using Gastric Alimetry were included. All were under the care of specialist gastroenterologists at a single tertiary referral centre in New Zealand (Waitematā, Auckland, New Zealand) between March 2019 and March 2022. Patients were excluded if they had a diagnosis of inflammatory bowel disease.

### Data collection

Demographic information, comorbidities such as depression or anxiety disorders, and medication use were collected. Baseline symptom-severity and quality-of-life was completed with the Patient Assessment of Upper Gastrointestinal Disorders-Symptom Severity Index (PAGI-SYM), Gastroparesis Cardinal Symptom Index (GCSI), Patient Assessment of Upper Gastrointestinal Disorders-Quality of Life (PAGI-QOL), and the EuroQol Visual Analogue Scale (EQ-VAS) instruments. ^19–21^

Beginning at the patient’s index presentation to the tertiary hospital for symptoms of CNVS, data on the type and number of investigations (upper gastrointestinal endoscopy, colonoscopy, ultrasound (USS) abdominal computed tomography (CT), abdominal magnetic resonance imaging (MRI), abdominal X-rays, gastric emptying scintigraphy, barium studies, oesophageal manometry and pH impedance studies), hospital admissions, emergency department (ED) presentations for gastroenterology issues, and outpatient gastroenterology appointments were recorded. To determine if the cause of admission or ED presentation was for a gastrointestinal problem, clinical note review was completed by two independent researchers with discrepancies resolved by a third. Duration of follow up was defined as the time between the index appointment and most recent encounter. Relevant gastrointestinal abnormalities that determined diagnostic yield included structural abnormalities identified on gastroscopy (oesophagitis, duodenitis, gastric ulcers, hiatal hernias), or abnormalities identified on radiological or clinical reports of gastric emptying scintigraphy, manometry, pH impedance testing and other tests which could reasonably explain the presenting complaint(s) of nausea and/or vomiting. In cases of repeat testing, only the first investigation per patient was used.

### Alimetry classification

Gastric Alimetry spectral analysis was evaluated according to the following major phenotypes: ^22^

i. *Gastric neuromuscular dysfunction:* Gastric rhythm disorder or low amplitude, indicated by Gastric Alimetry Rhythm Index (GA-RI) <0.25, and/or BMI-adjusted amplitude <22 μV.^13^ These patients were managed as per gastroparesis guidelines,^23^ regardless of GET status.
ii. *High sustained BMI-adjusted amplitude*: >70 μV.^13^ Gastric outlet resistance was considered if gastric emptying was also delayed.^24^
iii. *High frequency phenotype:* >3.35cpm. Vagal dysfunction was considered, particularly if concomitant diabetes or prior oesophagogastric surgery.^14,25^

Gastric Alimetry Symptom Profiling was evaluated according to the following major phenotypes:

i. *Sensorimotor:* symptoms correlating with gastric amplitude, suggesting a likely hypersensitivity / accommodation disorder.^26^
ii. *Post-gastric*: symptoms trending upward late in post-prandial period; after the gastric meal response had peaked, indicating a likely more distal pathologies (e.g. small bowel dysmotility or other disorders).^27^
iii. *Continuous profile*: symptoms constant / do not correlate with gastric amplitude. In these patients, gut-brain axis disorders were considered if spectral analysis was normal, per the study of Gharibans et al.^28^

For this study, a spectral abnormality or continuous or sensorimotor symptom profile identified via Gastric Alimetry was considered a positive diagnostic finding.

### Healthcare utilisation

The primary outcome of this study was healthcare utilisation and associated costs of patients, arising from gastrointestinal investigations, emergency room presentations, gastroenterology clinic visits and hospital admissions. Costs of investigations and healthcare visits were supplied from New Zealand Hospital administrators Pharmac (a centralised government procurement agency), hospital costing data and radiology centres sourced from data relevant to 2023. Where a range of prices were available for a procedure, the average price was used (**Table S1**).

Descriptive analysis of healthcare utilisation costs was completed, with data presented as frequency and percentages or median and interquartile ranges unless otherwise specified. Annual cost of care, and relative contributions from different cost sources were calculated. Average healthcare utilisation costs per patient were calculated before vs after Gastric Alimetry testing. Follow-up was adjusted for follow up duration, and pre- and post-comparisons were compared using non-parametric Wilcoxon paired analysis. All analyses were performed in R version 4.0.3 (R Foundation for Statistical Computing, Vienna, Austria).

### Health economic analysis

A decision tree outlining the diagnostic pathway for patients with chronic nausea and vomiting syndromes (based on the Rome IV diagnostic algorithms for gastroduodenal disorders and UEG consensus statements,^29,30^), with and without the incorporation of Gastric Alimetry, is presented in **Figure 1**. A linear pathway to investigations was assumed to enable the analysis, accepting that in practice, the choice and sequence of investigations is tailored to each individual patient.

**Figure 1:**
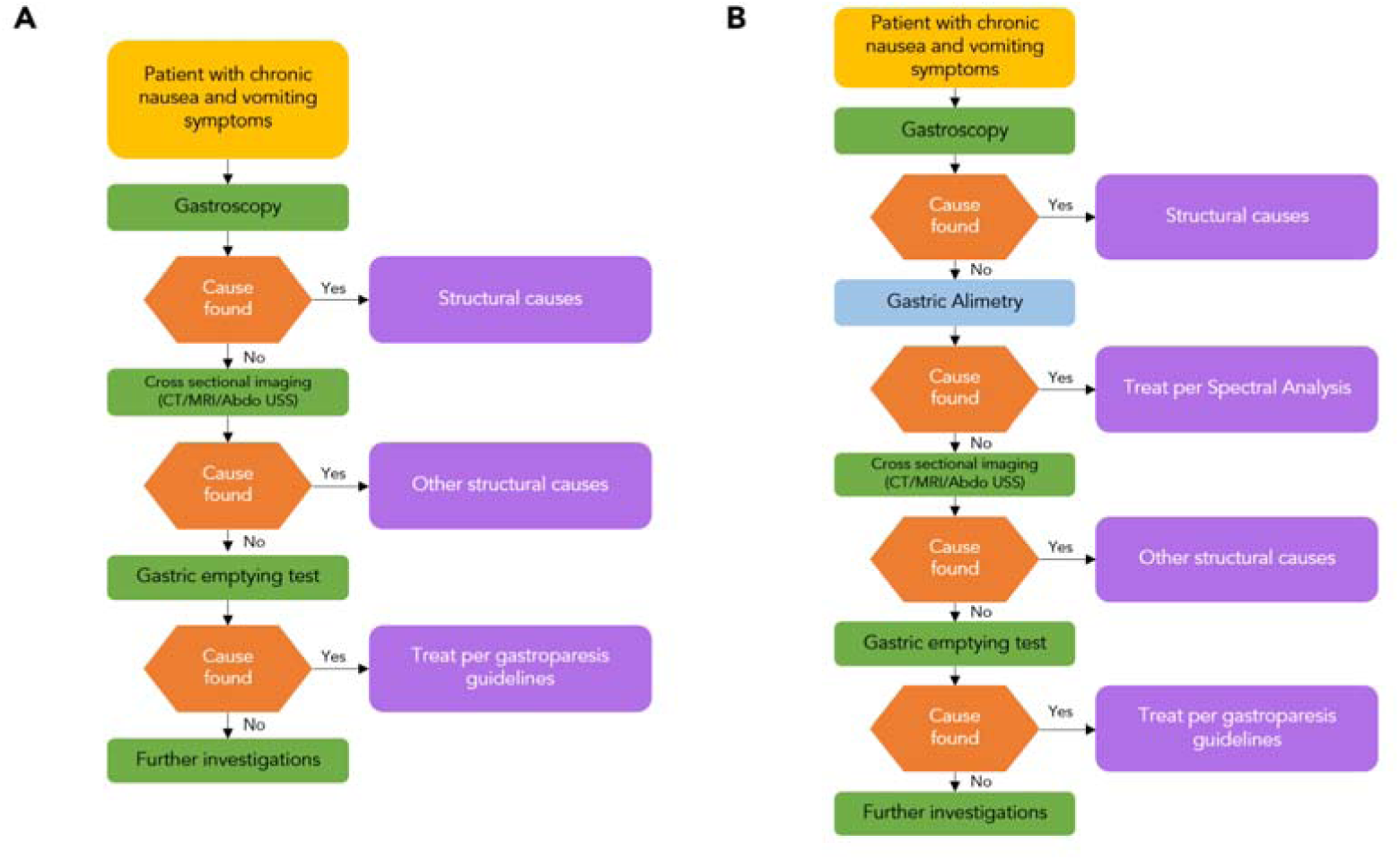
Flowchart of investigations. Model of investigations in NVS, based on literature and the evaluated clinical cohort. A) Standard care and B) incorporating Gastric Alimetry into investigations. A linear pathway to investigations was assumed to enable the analysis, accepting that in practice, the choice and sequence of investigations is tailored to each individual patient.

Endoscopy, radiology, and gastric emptying testing were included as the decision nodes for the decision tree analysis as these tests have the greatest weight in informing further investigation and treatment decision making and each represent significant healthcare expenditure. The standard care pathway was informed by the Rome IV diagnostic algorithms for gastroduodenal disorders and UEG consensus statements,^29,30^ and by prior qualitative work.^11^ In both pathways patients require gastroscopy initially to ensure no structural abnormalities. In the comparative pathway patients undergo Gastric Alimetry prior to radiological investigations as this is a less invasive test. Less expensive procedures such as blood tests and X-ray were not included in the model due to vast heterogeneity in the utilisation of these healthcare resources. Indirect costs to patients, pharmaceuticals, and nutritional support costs were not included.

The model probabilities were populated using diagnostic yield for each investigation (number of tests identifying an abnormality divided by the total number of tests performed) in the present patient cohort. A sensitivity analysis was conducted by populating the decision tree with the probabilities based upon diagnostic yield from an American gastroparesis cohort study.^7^ Costs of investigations and healthcare visits were supplied from New Zealand Hospital administrators sourced from data relevant to 2023 as described above. An overall cost for cross sectional imaging was estimated based on CT scan costs (**Table S1**). The cost of further tests if no abnormality was found was set at an additional estimated $2000 NZD based on the additional costs of investigations including esophageal manometry, pH impedance, lower gastrointestinal endoscopy, and barium swallow studies (**Table S1**).

## Results

### Patient demographics

Overall, 31 consecutive patients with symptoms of nausea, vomiting, abdominal pain, and bloating under the care of a specialist gastroenterologist were evaluated with Gastric Alimetry from March 2019 to March 2022. Characteristics are detailed in **Table 1**, including age, sex, BMI, presenting symptoms and comorbidities. The mean age was 36.1 years (range 16 to 66) with a female preponderance (26/31; 83.9%). The most commonly reported presenting symptoms were nausea (83.9%), abdominal pain (61.3%), vomiting (67.7%) and bloating (35.5%). There was a significant burden of gastrointestinal symptoms in this cohort with an overall median PAGI-SYM score of patients of 2.9 out of 5 (IQR 2.4 to 3.2) and a median GCSI score of 3.13 out of 5 (IQR 2.6 to 3.6). Patients had a median PAGI-QoL score of 2.7 out of 5 (IQR 2.2 to 3.2). Median EQ-VAS quality of life score was 59.0 out of 100 (IQR: 31.5 to 72.0; 0 is the worst possible and 100 is the best possible).

**Table 1:** Baseline characteristics.

| Variable | n = 31 |
| --- | --- |
| Age | 36.1 +/- 15.8 |
| Sex (female) | 26 (83.9%) |
| BMI | 23.9 +/- 5.4 |
| Ethnicity |  |
| NZ European | 22/31 (71.0%) |
| Māori | 2/31 (6.5%) |
| Other | 7/31 (22.5%) |
| Rome Criteria |  |
| CNVS and FD | 29/31 (93.5%) |
| FD alone | 2/31 (6.5%) |
| Presenting symptoms at index presentation |  |
| Nausea | 26 (83.9%) |
| Vomiting | 19 (61.3%) |
| Abdominal pain | 19 (67.7%) |
| Weight loss | 8 (25.8%) |
| Stomach burn/heartburn | 4 (12.9%) |
| Bloating | 11 (35.5%) |
| Early fullness and satiety | 3 (9.7%) |
| Postprandial fullness | 4 (12.9%) |
| Comorbid anxiety and depression |  |
| Anxiety | 12 (38.7%) |
| Depression | 6 (19.4%) |

Psychological comorbidities were common; 12 patients had a coexisting anxiety disorder (38.7%) and 6 a coexisting major depressive disorder (19.4). Four patients had diabetes mellitus (12.9%), and 5 patients had previous thorax or abdominal surgery (16.1%); heart and lung transplant, excision of endometriosis, high anterior resection, fundoplication, and laparoscopic duodeno-jejunostomy. No patients had a diagnosis of an eating disorder.

### Gastric Alimetry Testing

On average, Gastric Alimetry testing was completed 2.8 years from index interaction with the healthcare system (range 0.1 to 15.8 years). Gastric Alimetry spectral abnormalities were found on 14 tests (45.2%); including sustained dysrhythmia indicating likely neuromuscular dysfunction (7), high gastric frequencies (4), and high gastric amplitude (3). Gastric Alimetry symptom profiling identified 7 patients (22.6%) with a continuous symptom phenotype in the presence of a normal spectral analysis, considered to indicate a possible disorder of the gut-brain axis.^28^ An additional 2 patients (6.5%) had sensorimotor symptom profiles. In total Gastric Alimetry had a positive finding in 23 patients (74.2%).

### Healthcare utilisation

Median follow up time per patient from index presentation was 4.7 years (range: 1.3 to 17.2 years). Over 148 patient-years of follow-up, there were a total of 215 emergency department presentations, 95 hospital admissions, 147 gastroenterology consultations, 39 gastroscopies, 11 barium swallows, 14 colonoscopies, 21 ultrasounds, 24 abdominal CT, 8 MR enterography, 17 gastric emptying scintigraphy tests, 6 esophageal manometry tests, 5 pH impedance tests (**Figure 3**). An additional 67 gastroscopies were performed for feeding tube placement (but have not been included in the cost evaluation as this was a therapeutic procedure). 22 patients presented to ED at least once (71.0%), and 11 patients had at least one hospital admission greater than 24 hours (35.5%). Healthcare utilisation showed substantial heterogeneity, with 6 patients (19.4%) accounting for 80% of emergency room visits, 91.6% of patient admissions, and 29.2% of gastroenterology visits. Healthcare events remained substantial over time; the average number of all healthcare presentations per year was 1.4 events in year one, 0.7 in year 2, 0.4 in year 3, 0.8 in year 4, and 0.9 events in year five (p = 0.999; **Figure 2**).

**Figure 2:**
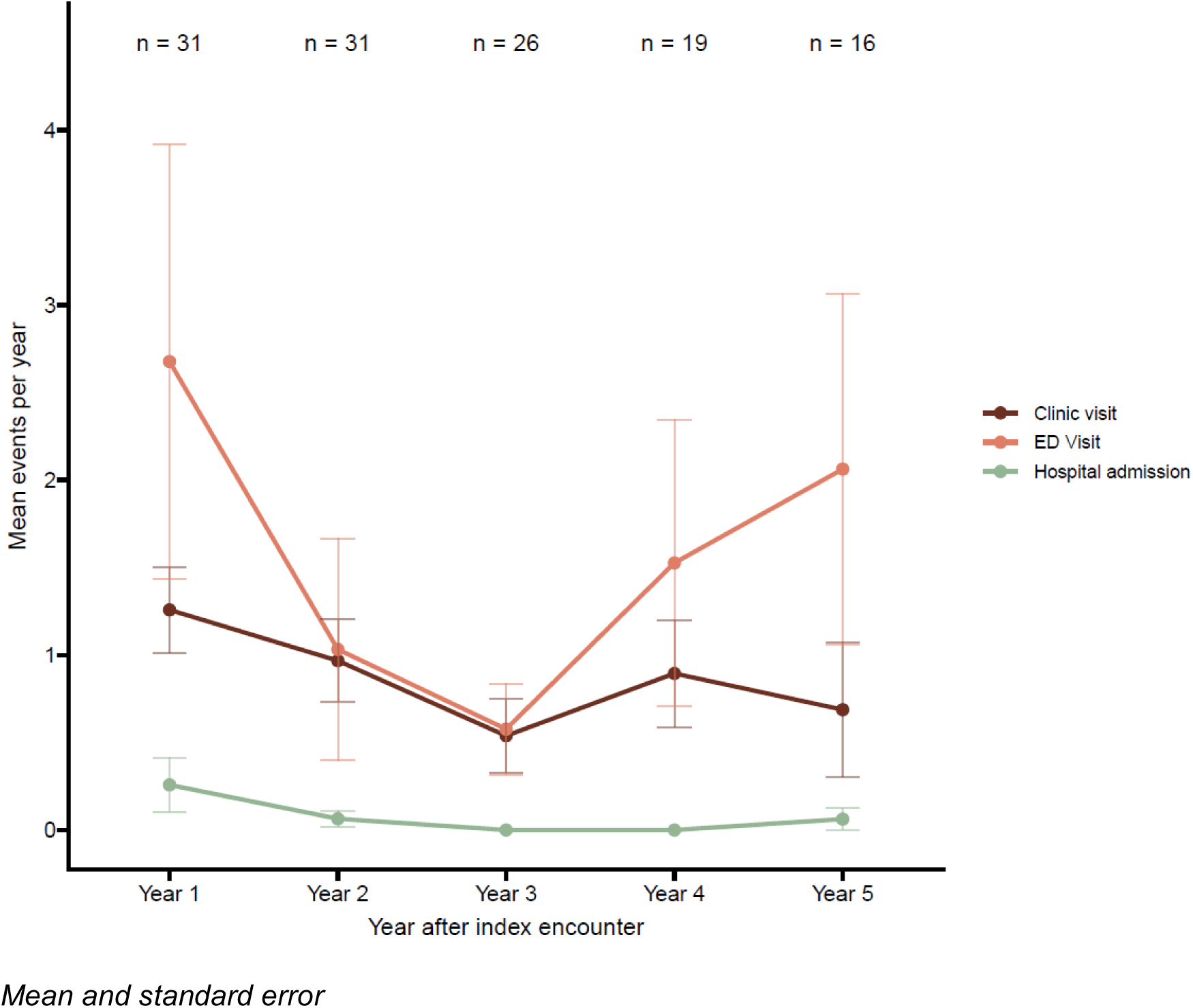
Healthcare events from index presentation per patient-year of follow up: gastroenterology consultations, emergency department presentations and inpatient admissions.

**Figure 3:**
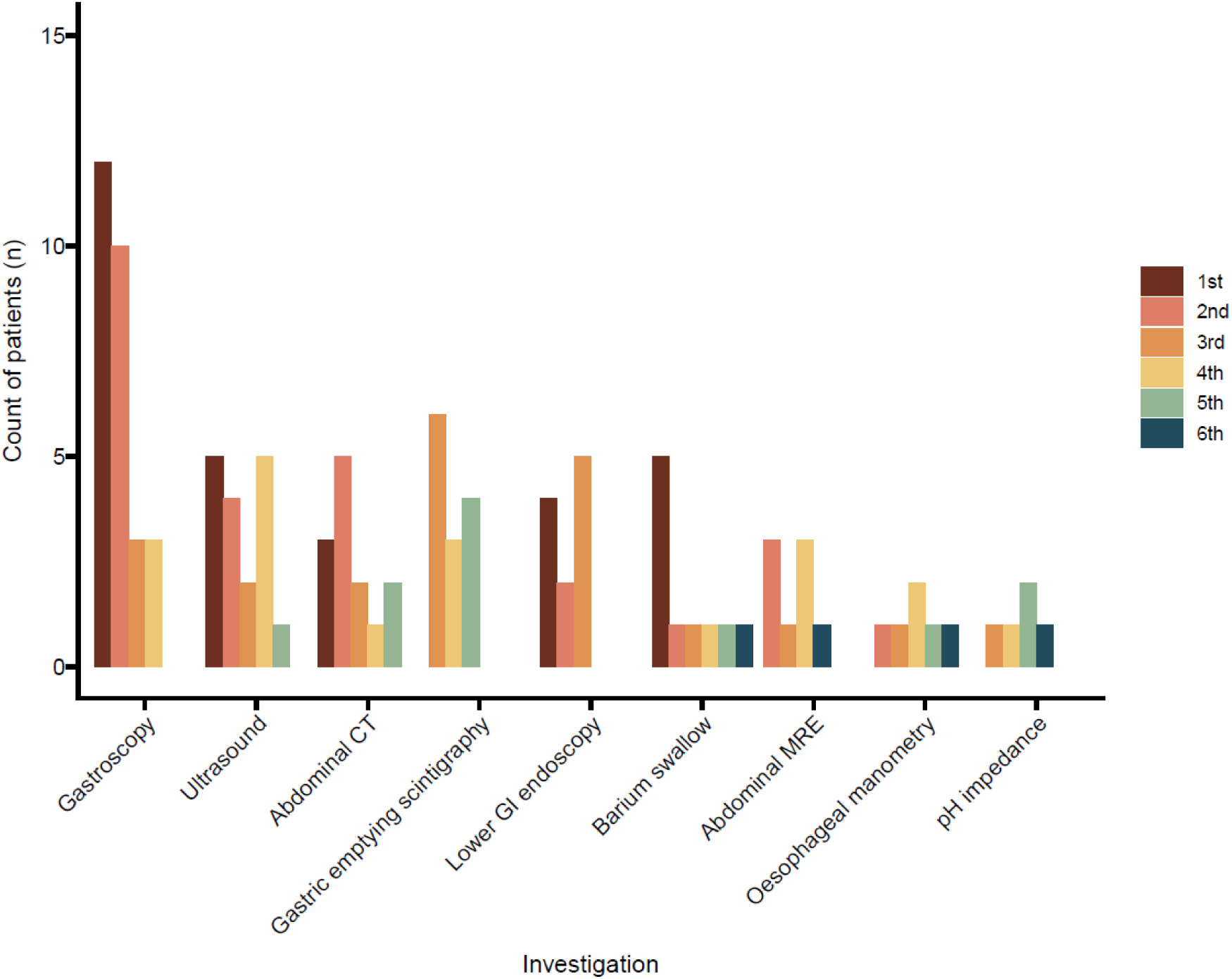
Investigations performed as part of clinical care. Legend refers to the order that investigation was performed in (1st: first line index investigation).

Overall investigations undertaken are presented in **Figure 3** and the diagnostic yield of the investigations is presented in **Table 2**. 39 gastroscopies were conducted in the public healthcare system in 28 patients. Various pathology was found during the gastroscopies (9/39, 23.1%); coeliac disease (1), duodenitis (1), esophagitis (1), small hiatal hernia or gastroesophageal laxity (4). Eight of 28 patients (28.6%) underwent repeat diagnostic gastroscopies with none identifying further abnormalities. 53 cross-sectional imaging studies (USS, CT, MRE) were undertaken in 23 patients (74.2%); 11 patients underwent abdominal USS, 13 patients had CT scans, and 8 patients had MRE. Only 7 cross-sectional imaging studies found abnormalities (13.2%); USS identified cholecystitis in 1 patient (9.1%); four CT scans identified relevant gastrointestinal abnormalities (16.7%; cholecystitis, SMA syndrome, epiploic appendagitis, annular pancreas), MRE identified abnormalities in 2 patients (25%; large bowel malrotation, slow bowel transit). 11 patients underwent repeat CT after a prior scan had not detected an abnormality; all had no abnormalities detected on repeat imaging.

**Table 2:**
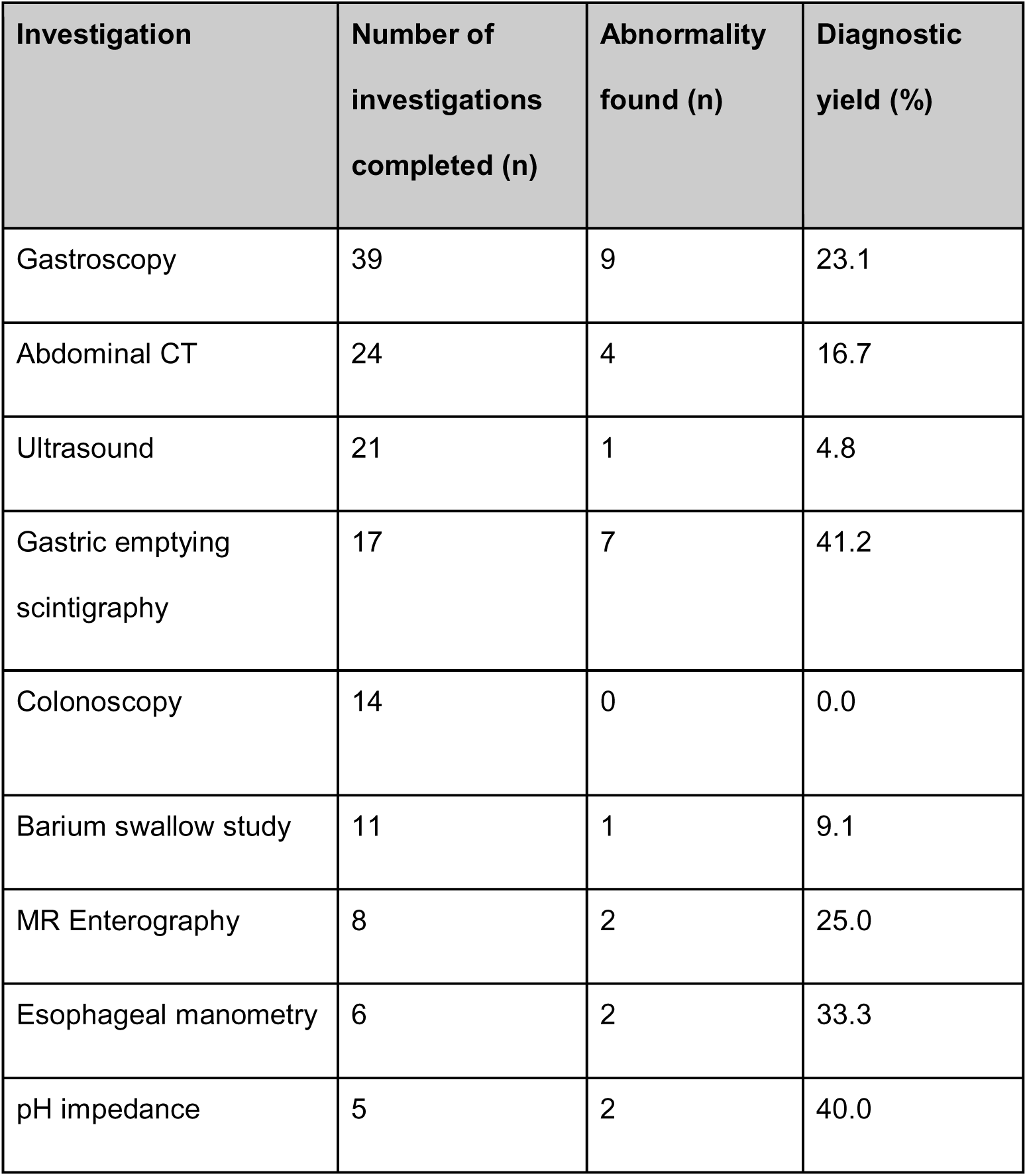
Diagnostic yield of conventional investigations. Table includes excludes testing within the same patient. CT: computerised tomography. MRE: magnetic resonance enterography

| Investigation | Number of investigations completed (n) | Abnormality found (n) | Diagnostic yield (%) |
| --- | --- | --- | --- |
| Gastroscopy | 39 | 9 | 23.1 |
| Abdominal CT | 24 | 4 | 16.7 |
| Ultrasound | 21 | 1 | 4.8 |
| Gastric emptying scintigraphy | 17 | 7 | 41.2 |
| Colonoscopy | 14 | 0 | 0.0 |
| Barium swallow study | 11 | 1 | 9.1 |
| MR Enterography | 8 | 2 | 25.0 |
| Esophageal manometry | 6 | 2 | 33.3 |
| pH impedance | 5 | 2 | 40.0 |

Gastric emptying scintigraphy was undertaken 17 times in 15 patients and demonstrated delayed gastric emptying in 5/15 patients (33.3%) and accelerated gastric emptying in 1 patient (6.7%), identifying abnormalities in 40% of patients. In the 2 patients (13.3%) in whom, gastric emptying testing was repeated; one patient had consistent results of delayed gastric emptying, while the another showed normal gastric emptying after a previously delayed emptying test result.

### Before and after Alimetry

In the current clinical cohort, the costs of all healthcare utilisation before and after Alimetry, adjusted for available follow up time is presented in **Figure 4**. The number of investigations, ED visits, clinic visits, and inpatient stays per year reduced substantially after Gastric Alimetry testing, compared to pre-testing healthcare utilisation (median $12,172 NZD, IQR $6,301 to $27,263 versus $839 NZD, IQR $263 to $2,520; p<0.001; **Figure 4A**). This finding remained consistent when only considering the cost of ED visits, clinic visits, and inpatient hospitalisations to adjust for higher investigation costs early in the natural history of disease (median $4,366 NZD, IQR $1,823 to $15,374 versus $504 NZD, IQR $134 to $1,466; **Figure 4B**). Associated cost savings per patient-year in overall healthcare utilisation including investigations were $12,032 NZD (IQR $2,746 to $26,108). Associated cost savings per patient-year in healthcare visits alone were $3,512 NZD (IQR $1,034 to $14,706).

**Figure 4:**
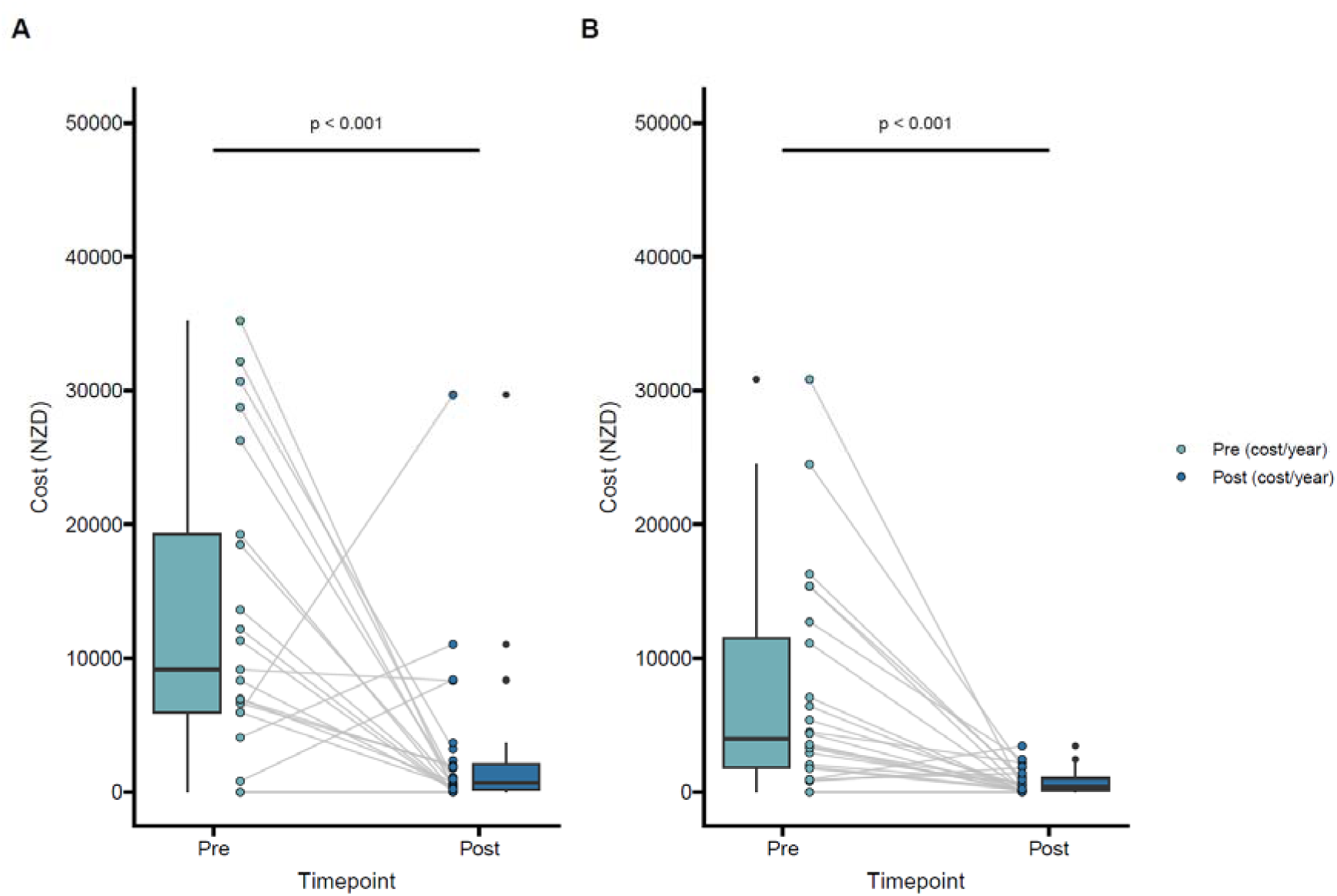
Costs of healthcare utilisation before and after Alimetry testing. A: Total healthcare utilisation costs including costs of investigations as well as emergency department (ED) visits, clinic visits and inpatient medical stays. B: Costs of ED visits, clinic visits and inpatient medical stays only excluding costs of investigations.

### Decision tree analysis for clinical investigations

The decision tree was used to estimate the costs of a standard care pathway versus a care pathway incorporating Gastric Alimetry after gastroscopy (**Table 3**; **Figure 5**). The diagnostic yields for the clinical investigations from the prior section were used to populate the probabilities of moving between nodes in the decision tree. The expected cost of the standard care diagnostic pathway was $4,610.53 and the expected cost of a diagnostic pathway incorporating Gastric Alimetry after normal gastroscopy was $2,977.93. A sensitivity analysis was undertaken where the decision tree probabilities were populated from diagnostic yields reported by Dudekula et al. from a large US cohort of comparable patients (**Table 3**) ^7^. On sensitivity analysis, expected costs of standard diagnostic pathway was $3,893.31 per patient versus $2,774.07 when incorporating Gastric Alimetry.

**Figure 5:**
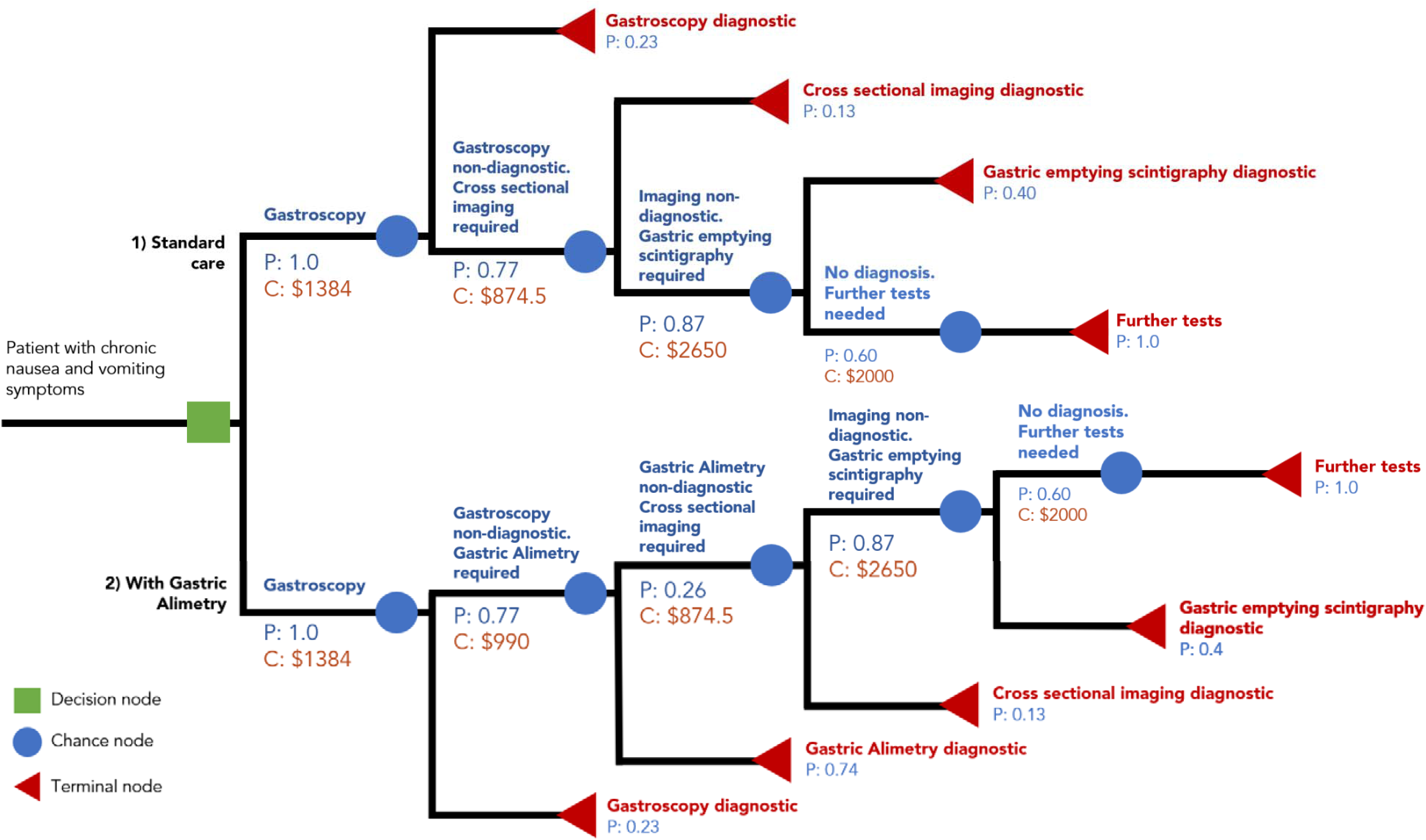
Decision tree analysis of standard care vs incorporation of Gastric Alimetry.

**Table 3:** Estimated costs using standard care vs Gastric Alimetry pathway.

| Using local diagnostic values | Standard Care |  | Gastric Alimetry Pathway |  |
| --- | --- | --- | --- | --- |
| Terminal node outcome | Probability of flowchart outcome | Cost | Probability of flowchart outcome | Cost |
| Gastroscopy diagnostic | 0.23 | 1384.23 | 0.23 | 1384.23 |
| Gastroscopy is non-diagnostic, Gastric Alimetry diagnostic | | | 0.57 | \$2,374.23 |
| Gastroscopy, and Gastric Alimetry are non-diagnostic but cross-sectional imaging diagnostic | 0.10 | \$2,258.71 | 0.03 | \$3,248.71 |
| Gastroscopy, Gastric Alimetry and cross-section imaging are non-diagnostic but GES diagnostic | 0.28 | \$4,908.71 | 0.07 | \$5,898.71 |
| All prior tests are non-diagnostic, further tests needed | 0.39 | \$6,908.71 | 0.10 | \$7,898.71 |
| Expected values | \$4,610.53 | | \$2,977.93 | |

| Using Dudekula et al<br>diagnostic values | Standard Care |  | Gastric Alimetry Pathway |  |
| --- | --- | --- | --- | --- |
| Terminal node outcome | Probability<br>of<br>flowchart<br>outcome | Cost | Probability<br>of<br>flowchart<br>outcome | Cost |
| Gastroscopy diagnostic | 0.25 | 1384.23 | 0.25 | 1384.23 |
| Gastroscopy is non-diagnostic,<br>Gastric Alimetry diagnostic | | | 0.56 | \$2,374.23 |
| Gastroscopy, and Gastric<br>Alimetry are non-diagnostic<br>but cross-sectional imaging<br>diagnostic | 0.10 | \$2,258.71 | 0.03 | \$3,248.71 |
| Gastroscopy, Gastric Alimetry<br>and cross-section imaging are<br>non-diagnostic but GES<br>diagnostic | 0.59 | \$4,908.71 | 0.15 | \$5,898.71 |
| All prior tests are non-diagnostic, further tests<br>needed | 0.06 | \$6,908.71 | 0.02 | \$7,898.71 |
| Expected values | \$3,893.31 | | \$2,774.07 | |

## Discussion

This study aimed to define healthcare utilisation among patients with NVS in a tertiary care setting, to evaluate the diagnostic yield of associated investigations, and to estimate the potential health economic impact of implementing BSGM testing using the Gastric Alimetry system using both real-world data and a decision-tree analysis. The real-world data confirmed that NVS care constitutes a substantial healthcare burden, characterised by repeated healthcare visits and high rates of investigations. The introduction of Gastric Alimetry was associated with a substantial reduction in healthcare costs (>$10,000 NZD per patient), due principally to reduced investigations, acute care presentations, and inpatient stays. The decision-tree analysis supported these findings, estimating savings of >30% with regard to diagnostic investigations alone. This study therefore demonstrates the potential utility of Gastric Alimetry to reduce healthcare costs by improving diagnostic clarity.

The high rates of healthcare utilisation reported in this New Zealand study were consistent with other data, including studies from the US healthcare context.^7^ Several studies have shown high rates of diagnostic testing, acute care presentations and inpatient admissions in NVS patients, which are sustained over multiple years.^7,8,17,31^ Investigations are often repeated in a confirmatory fashion, as again shown in the current study, with repeat gastroscopy and abdominal CT rates of 29% and 85% respectively. Patients may therefore also exceed annual recommended radiation limits due to repeated medical imaging.^7^ This ongoing testing occurs despite clinical recommendations in functional disorders,^29^ and is explained by the clinical uncertainty commonly experienced when facing distressing gastric symptoms that fluctuate in intensity. Our data again show that such repeated testing is rarely beneficial in patients with chronic gastric symptoms, but also indicate that Gastric Alimetry may act as a ‘circuit-breaker’ in this diagnostic process through a positive diagnostic yield.

Based on previous literature, US healthcare costs for NVS are expected to be higher than those described in this study.^9,32,33^ For example, a recent detailed analysis by Chen et al, found that in gastroparesis US healthcare costs ranged from US$34,885 to $14,396 in diabetic and idiopathic disease over the first three years of diagnosis.^8^ Differences in cost may be in part due to the public healthcare funding model in the New Zealand context in which this study was performed. The benefits defined by this study may therefore be potentially amplified in the US setting, where costs of care are higher.

The results of this study indirectly support the capability of Gastric Alimetry to provide improved diagnostic accuracy in a manner that informs more targeted therapy. This was also supported by a separate study by Varghese et al, in which it was shown that Gastric Alimetry aided management decisions in >80% of patients.^17^ One key advantage of this modality, as opposed to gastric emptying, is to identify underlying neuromuscular disorders, which are well established to occur in NVS, but which have previously been difficult to separate without full thickness tissue biopsies.^5,28,34,35^ In addition, the combination of spectral and symptom-based phenotypes cover a broader range of diagnostic possibilities, including gut-brain axis disorders, sensorimotor disorders, and the sequelae of long-term diabetes.^14,15,18^ Therefore, as demonstrated in the present decision-tree analysis, the incorporation of the test earlier in the diagnostic pathway may decrease the ongoing need for negative confirmatory diagnostic testing and trial-and-error therapy. Additional prospective and international data is now awaited to confirm and extend these findings in additional cohorts and care settings.

A limitation of this study should also be noted that pharmaceutical and nutrition support costs were not included, as it was found these costs could not be as reliably computed in a retrospective data analysis. Nutrition costs may be particularly significant, given other recent studies showing that Gastric Alimetry may aid in the selection of patients for invasive nutrition,^16,17^ by more robustly defining patient subgroups with true gastrointestinal neuromuscular disorders from those with gut-brain axis or sensorimotor disorders.^18,28^ Given the high costs of nutrition support, which may extend to hundreds of thousands of dollars per year for patients on parenteral nutrition,^36^ the cost savings identified in this study could therefore have been underestimated. For example, Varghese et al recently reported higher average annualised cost savings of NZ$19,787 following the introduction of Gastric Alimetry in a cohort of patients in whom enteral support costs were able to be defined, and which included patients on parenteral nutrition.^17^ This study also indicated that Gastric Alimetry may aid in the de-escalation of pharmacological therapy in approximately 20% of cases, potentially contributing additional savings.^17^

The reported data are likely to be reliable because healthcare utilisation data can be robustly retrieved, and cases were captured consecutively, and although private care episodes and general practice visits were not captured these were expected to be infrequent in the study population context. In addition, patient out-of-pocket expenses may be substantial in functional gastroduodenal disorders,^9^ but were not a focus of the current study. The retrospective nature of the study also meant that patient outcomes and quality of life were not assessed, as these endpoints generally require validated questionnaires that are best conducted prospectively. Pragmatic assumptions based on recent clinical guidelines were made with regards to the linear diagnostic pathways employed in the decision tree analysis, whereas in practice physicians choose investigations as tailored to a specific presentation and patient context. It should also be noted that the diagnostic journey is not a linear pathway for these patients. Multiple interactions occur with the healthcare system prior to diagnosis (including blood tests, x-rays, consultations, emergency department presentations which have not been incorporated), such that a simplified decision tree analysis was necessary to enable meaningful comparisons. Nevertheless, the linear pathway model was a reasonable assumption as it matches general guidelines for patient care pathways.

The real-world data on health economic outcomes performed on an annualised before vs after basis could have been confounded by a naturally decreasing intensity of investigations and presentations over the course of the disorder. However, previous studies have shown that NVS patients generally continue to be high healthcare users, with relatively modest reductions in care intensity over time,^7,8^ such that the sharp and substantial reductions in overall healthcare utilisation reported here were likely to be meaningful. In addition, it was recognized that healthcare utilisation changes were skewed, with a minority of patients contributing most to the cost reductions, which is a common feature of health economics studies. Future studies should continue to evaluate the management changes, healthcare outcomes, and health economics associated with the introduction of Gastric Alimetry as usage of the test expands internationally, including in prospective studies.

In conclusion, this study presents real-world data together with a decision tree analysis of diagnostic workflows, which show that Gastric Alimetry may improve the clinical care pathway in NVS, resulting in reductions in healthcare utilisation and cost. Non-contributory or confirmatory investigations remain common practice in NVS management, contributing to iatrogenic harm through radiation exposure, but this problem can be mitigated by streamlined diagnostic pathways.

## Author Contributions

WX, LW, CV, CC, CD, CK were involved in study conception and design. WX, LW, CV, CK were involved in data collection. All authors were involved in the data analysis and interpretation, drafting, critical revisions, and final approval of the manuscript.

## Data Availability Statement

The data that support the findings of this study are available from the corresponding author upon reasonable request.

## Supplementary material

**Table S1:** Costs of investigations. Costs were calculated during January 2023 in NZD

| Procedure | Cost (NZD) |
| --- | --- |
| ED visit: duration 0-2 Hours | \$400 |
| ED visit: duration 2-23 hours | \$750 |
| ED stay with transfer to inpatient | \$1,300 |
| Medical Inpatient (per 24 hour stay) | \$1,200 |
| Medical Inpatient (Day stay) | \$600 |
| Gastroenterology clinical appointment (Initial) | \$675.85 |
| Gastroenterology clinical appt (Follow Up) | \$385.78 |
| Gastroscopy | \$1,384.23 |
| Ultrasound | 298.96 |
| Abdominal CT | 874.48 |
| Colonoscopy | 2075.53 |
| Abdominal X-ray | 157.25 |
| MR enterography | 1686.25 |
| Gastric emptying scintigraphy | 2650 |
| Oesophageal manometry | 1063.49 |
| pH impedance | 1063.49 |
| Barium Study | 651.67 |
| Alimetry | 990 |

## Acknowledgements

We thank the staff and patients who participated in this study.

## References

1. Stanghellini V, Chan FKL, Hasler WL, Malagelada JR, Suzuki H, Tack J, et al. Gastroduodenal Disorders. Gastroenterology. 2016 May;150(6):1380–92.

2. Sperber AD, Bangdiwala SI, Drossman DA, Ghoshal UC, Simren M, Tack J, et al. Worldwide Prevalence and Burden of Functional Gastrointestinal Disorders, Results of Rome Foundation Global Study. Gastroenterology. 2021 Jan;160(1):99–114.e3.

3. Janssen P, Harris MS, Jones M, Masaoka T, Farré R, Törnblom H, et al. The relation between symptom improvement and gastric emptying in the treatment of diabetic and idiopathic gastroparesis. Am J Gastroenterol. 2013 Sep;108(9):1382–91.

4. Vijayvargiya P, Jameie-Oskooei S, Camilleri M, Chedid V, Erwin PJ, Murad MH. Association between delayed gastric emptying and upper gastrointestinal symptoms: a systematic review and meta-analysis. Gut. 2019 May;68(5):804–13.

5. Pasricha PJ, Grover M, Yates KP, Abell TL, Bernard CE, Koch KL, et al. Functional Dyspepsia and Gastroparesis in Tertiary Care are Interchangeable Syndromes With Common Clinical and Pathologic Features. Gastroenterology. 2021 May;160(6):2006– 17.

6. Carson DA, Bhat S, Hayes TCL, Gharibans AA, Andrews CN, O’Grady G, et al. Abnormalities on Electrogastrography in Nausea and Vomiting Syndromes: A Systematic Review, Meta-Analysis, and Comparison to Other Gastric Disorders. Dig Dis Sci [Internet]. 2021 May 6; Available from: 10.1007/s10620-021-07026-x

7. Dudekula A, O’Connell M, Bielefeldt K. Hospitalizations and testing in gastroparesis. J Gastroenterol Hepatol. 2011 Aug;26(8):1275–82.

8. Chen YJ, Tang W, Ionescu-Ittu R, Ayyagari R, Wu E, Huh SY, et al. Health-care resource use and costs associated with diabetic and idiopathic gastroparesis: A claims analysis of the first 3 years following the diagnosis of gastroparesis. Neurogastroenterol Motil. 2022 Mar 30;e14366.

9. Lacy BE, Crowell MD, Mathis C, Bauer D, Heinberg LJ. Gastroparesis: Quality of Life and Health Care Utilization. J Clin Gastroenterol. 2018 Jan;52(1):20–4.

10. Pavurala RB, Stanich PP, Krishna SG, Guturu P, Hinton A, Conwell DL, et al. Predictors of Early Readmissions in Hospitalized Patients With Gastroparesis: A Nationwide Analysis. J Neurogastroenterol Motil. 2021 Jul 30;27(3):408–18.

11. Sebaratnam G, Law M, Broadbent E, Gharibans A, Andrews C, Daker C, et al. A qualitative study of patient and clinician perspectives of the clinical care pathway for nausea and vomiting syndromes. Front Psychol [Internet]. 2023;14. Available from: https://www.frontiersin.org/articles/10.3389/fpsyg.2023.1232871

12. Gharibans A, Hayes T, Carson D, Calder S, Varghese C, Du P, et al. A novel scalable electrode array and system for non-invasively assessing gastric function using flexible electronics. Neurogastroenterology & Motility. 2022 Jun 14;e14418.

13. Varghese C, Schamberg G, Calder S, Waite S, Carson DA, Foong D, et al. Normative values for body surface gastric mapping evaluations of gastric motility using Gastric Alimetry: spectral analysis. The American Journal of Gastroenterology. 2023;118(6):1047–57.

14. Xu W, Gharibans AA, Calder S, Schamberg G, Walters A, Jang J, et al. Defining and phenotyping gastric abnormalities in long-term type 1 diabetes using body surface gastric mapping. Gastro Hep Advances [Internet]. 2023;In press. Available from: 10.1101/2022.08.10.22278649

15. Carson DA, O’Grady G, Du P, Gharibans AA, Andrews CN. Body surface mapping of the stomach: New directions for clinically evaluating gastric electrical activity. Neurogastroenterol Motil. 2021 Mar;33(3):e14048.

16. Varghese C, Xu W, Daker C, Bissett IP, Cederwall C. Clinical utility of Gastric Alimetry® in the management of intestinal failure patients with possible underlying gut motility disorders. Clin Nutr Open Sci [Internet]. 2023 Aug; Available from: http://www.clinicalnutritionopenscience.com/article/S2667268523000359/abstract

17. Daker C, Varghese C, Xu W, Cederwall C. Gastric Alimetry impacts the management pathway of chronic gastroduodenal disorders [Internet]. bioRxiv. 2023. Available from: https://www.medrxiv.org/content/10.1101/2023.02.06.23285567v1.abstract

18. Wang WJ, Foong D, Calder S, Schamberg G, Varghese C, Tack J, et al. Gastric Alimetry ® improves patient phenotyping in gastroduodenal disorders compared to gastric emptying scintigraphy alone. medRxiv [Internet]. 2023 May 25; Available from: 10.1101/2023.05.18.23290134

19. Rentz AM, Kahrilas P, Stanghellini V, Tack J, Talley NJ, De la loge C, et al. Development and psychometric evaluation of the patient assessment of upper gastrointestinal symptom severity index (PAGI-SYM) in patients with upper gastrointestinal disorders. Qual Life Res. 2004 Dec 1;13(10):1737–49.

20. De la loge C, Trudeau E, Marquis P, Kahrilas P, Stanghellini V, Talley NJ, et al. Cross-cultural development and validation of a patient self-administered questionnaire to assess quality of life in upper gastrointestinal disorders: The PAGI-QOL©. Qual Life Res. 2004 Dec 1;13(10):1751–62.

21. Herdman M, Gudex C, Lloyd A, Janssen M, Kind P, Parkin D, et al. Development and preliminary testing of the new five-level version of EQ-5D (EQ-5D-5L). Qual Life Res. 2011 Dec;20(10):1727–36.

22. O’Grady G, Varghese C, Schamberg G, Calder S, Du P, Xu W, et al. Principles and clinical methods of body surface gastric mapping: Technical review. Neurogastroenterol Motil. 2023 Mar 29;e14556.

23. Camilleri M, Kuo B, Nguyen L, Vaughn VM, Petrey J, Greer K, et al. ACG Clinical Guideline: Gastroparesis [Internet]. Vol. 117, American Journal of Gastroenterology. 2022. p. 1197–220. Available from: 10.14309/ajg.0000000000001874

24. Brzana RJ, Koch KL, Bingaman S. Gastric myoelectrical activity in patients with gastric outlet obstruction and idiopathic gastroparesis. Am J Gastroenterol. 1998 Oct;93(10):1803–9.

25. Carson DA, Robertson S, Wang THH, Varghese C, Gharibans AA, Windsor JA, et al. The Impact and Clinical Implications of Gastric Surgery on the Gastric Conduction System. Foregut. 2023 Mar 1;3(1):29–44.

26. Stanghellini V, Chan FKL, Hasler WL, Malagelada JR, Suzuki H, Tack J, et al. Gastroduodenal Disorders. Gastroenterology. 2016 May;150(6):1380–92.

27. Schamberg G, Varghese C, Uren E, Calder S, O’Grady G, Gharibans AA. Physiology-guided quantitative symptom analysis for gastroduodenal disorders [Internet]. bioRxiv. 2023. Available from: https://www.medrxiv.org/content/10.1101/2023.06.07.23291112v1.abstract

28. Gharibans AA, Calder S, Varghese C, Waite S, Schamberg G, Daker C, et al. Gastric dysfunction in patients with chronic nausea and vomiting syndromes defined by a noninvasive gastric mapping device. Sci Transl Med. 2022 Sep 21;14(663):eabq3544.

29. Tack J, Talley NJ. Gastroduodenal disorders. Am J Gastroenterol. 2010 Apr;105(4):757–63.

30. Schol J, Wauters L, Dickman R, Drug V, Mulak A, Serra J, et al. United European Gastroenterology (UEG) and European Society for Neurogastroenterology and Motility (ESNM) consensus on gastroparesis. United European Gastroenterol J. 2021 Apr;9(3):287–306.

31. Wang YR, Fisher RS, Parkman HP. Gastroparesis-related hospitalizations in the United States: trends, characteristics, and outcomes, 1995-2004. Am J Gastroenterol. 2008 Feb;103(2):313–22.

32. Wadhwa V, Mehta D, Jobanputra Y, Lopez R, Thota PN, Sanaka MR. Healthcare utilization and costs associated with gastroparesis. World J Gastroenterol. 2017 Jun 28;23(24):4428–36.

33. Hirsch W, Nee J, Ballou S, Petersen T, Friedlander D, Lee HN, et al. Emergency Department Burden of Gastroparesis in the United States, 2006 to 2013. J Clin Gastroenterol. 2019 Feb;53(2):109–13.

34. Grover M, Farrugia G, Lurken MS, Bernard CE, Faussone-Pellegrini MS, Smyrk TC, et al. Cellular changes in diabetic and idiopathic gastroparesis. Gastroenterology. 2011 May;140(5):1575–85.e8.

35. Angeli TR, Cheng LK, Du P, Wang THH, Bernard CE, Vannucchi MG, et al. Loss of Interstitial Cells of Cajal and Patterns of Gastric Dysrhythmia in Patients With Chronic Unexplained Nausea and Vomiting. Gastroenterology. 2015 Jul;149(1):56–66.e5.

36. Arhip L, Serrano-Moreno C, Romero I, Camblor M, Cuerda C. The economic costs of home parenteral nutrition: Systematic review of partial and full economic evaluations. Clin Nutr. 2021 Feb;40(2):339–49.

